# Strengthening evidence for text-based telehealth in post-operative care: A pragmatic study of the reach and effectiveness of two-way, text-based follow-up after voluntary medical male circumcision in South Africa

**DOI:** 10.1101/2024.12.05.24317906

**Authors:** Caryl Feldacker, Isabella Fabens, Tracy Dong, Khumbulani Moyo, Calsile Makhele, Motshana Phohole, Nelson Igaba, Sizwe Hlongwane, Jacqueline Pienaar, Maria Sardini, Felex Ndebele, Hannock Tweya, Marrianne Holec, Evelyn Waweru, Geoffrey Setswe

## Abstract

Building upon evidence of safety and efficiency gains from a randomized control trial (RCT) in South Africa, we further scaled implementation of two-way, short message service (SMS), text-based (2wT) follow-up after voluntary medical male circumcision (VMMC). We aimed to determine if gains in adverse event (AE) identification and reduced follow-up visits could be maintained when 2wT was implemented in routine VMMC settings. A pragmatic, stepped wedge design (SWD) study was implemented across three districts in South Africa. Men ages 15 and older could opt into the 2wT telehealth follow-up approach when their facility was in the intervention period. Men in routine periods were offered the standard of care (SoC): in-person post-operative visits on days 2 and 7 as per national VMMC guidelines. 2wT participants were not required to attend any postoperative visits but could return for care if desired or referred. Two quality of care markers, safety (AE ascertainment rate) and efficiency (# in-person follow-up visits), were compared between groups. We aimed for at least 200 men per step to have 80% power to detect a change in AE rate from before to after 2wT was implemented. Secondary analysis explored response rates; client and site uptake; and AE details. Among 6842 clients in the intervention period, 2856 opted into 2wT (37.8%) across three intervention waves and two platforms (SMS or WhatsApp). Among those with post-operative follow-up, the AE ascertainment rate was higher among 2wT (0.60%) than SoC (0.13%) clients (p = 0.0018), demonstrating safety gains. On average, 2wT participants had 2.1 fewer visits compared to SoC clients (p<0.001), demonstrating gains in follow-up efficiency. Among 2wT men, 2069/2586 (80%) responded via 2wT over 14 days, demonstrating engagement in post-operative care. Of all intervention clients, 93 2wT (3.6%) and 342 (8.0%) SoC were considered lost to follow-up. In this expansion trial, we provided additional evidence that the 2wT approach maintains the quality of post-operative care for adult VMMC clients. 2wT should be scaled to augment in-person, post-operative visits after VMMC for eligible, interested males ages 15 and older. To achieve potential impact, effort is needed to improve access and uptake to 2wT among providers and sites, expanding the 2wT approach for other acute follow-up care especially among men.

## Introduction

Voluntary medical male circumcision (VMMC) is a critical HIV prevention intervention with global support for sustained implementation across sub-Saharan Africa (SSA) (1, 2). VMMC is safe and major complications are rare (3); yet, many global guidelines still recommend at least one, in-person follow-up within 14 days (4). Our previous research in Zimbabwe and South Africa indicates that replacing mandatory in-person follow-up visits with two-way 2wT between patients and providers significantly improves the efficiency and safety of VMMC services. This approach reduces follow-up visits by 85% while doubling the identification of adverse events (AEs) (5–11). Expansion into routine VMMC service delivery in Zimbabwe suggests that 2wT brings other benefits alongside safety and efficiency gains (12, 13). To improve data quality, 2wT verifies unique VMMC client enrollments and facilitates compliance with follow-up care via documented messaging between clients and clinicians, reducing potential fraud of either enrollment or overreporting of post-operative visit attendance. 2wT also supports quality improvement initiatives by documenting longitudinal interactions between clients and clinicians, helping identify program weaknesses like gaps in post-operative counseling or wound care instructions that can trigger quality improvement efforts. 2wT also helps identify and document AEs earlier as compared to standard of care (SoC), likely reducing AE severity. Decision support tools, like alerts, for clinicians urge closing of referral loops when clients are asked to return for in-person reviews and prompts providers to follow-up with clients who do not respond, decreasing loss-to-follow-up (LTFU). This evidence is lacking in routine South Africa settings.

While digital or mobile health (mHealth) interventions are widely recognized for potential to improve healthcare quality, access, and costs (14–19), the striking potential benefits of 2wT are most evident in consideration of quality gaps and shrinking global HIV/AIDS budgets. 2wT at scale could vastly reduce unnecessary visits among males who are healing well while preserving client safety, safeguarding critical resources for optimizing quality VMMC service delivery. Balancing feasibility with the need for rigorous evidence to assess 2wT impact within routine VMMC programs outside of Zimbabwe, we conducted a quasi-experimental, opt-in, modified stepped wedge design (SWD) study in South Africa. The 2wT scale-up study in South Africa was implemented as part of routine VMMC follow-up in these sites with minimal oversight from an external, study-specific 2wT quality assurance team. We apply the concepts from the implementation science RE-AIM framework (Reach, Effectiveness, Adoption, Implementation, and Maintenance) to guide evaluation of 2wT implementation, applying findings for 2wT practice and policy considerations for expansion.

Our primary objectives in the 2wT SWD study were to examine 2wT reach (opt-in enrollment) and effectiveness (2wT safety and efficiency) at scale, comparing these outcomes between clients who opted into 2wT and those who remained in standard of care (SoC) with in-person reviews. As secondary outcomes, we explored 2wT client response rates; 2wT uptake among clients and sites; and detailed AE ascertainment across groups. We aimed to use findings to inform recommendations and strategies for increased 2wT reach and effectiveness at scale.

## Methods

### 2wT Technology Overview

The open-source 2wT system and intervention in both Zimbabwe and South Africa have been described in detail previously (5, 6, 12, 13). In brief, the 2wT telehealth approach provides an option for text-based follow-up for adult males for the critical 14 days after VMMC instead of required SoC, in-person, post-operative visits. For clients, 2wT requires only basic mobile phone or smartphone, to communicate via text with a VMMC nurse. 2wT combines automated educational messages on wound care with prompts on alternative days that request a response on healing (response 0=no problem; response 1= I have a concern). Messages are sent during the 14 days after the VMMC procedure; clients can interact with the VMMC nurse at any time after VMMC, including beyond the 14 days. Clients who responded with a 1 could engage directly with a 2wT VMMC nurse who triaged them to care, if needed, or provided information or reassurance by message or call, if requested. Men could choose to receive SMS messages via Afrikaans, English, isi-Zulu, Setswana and Sesotho, though WhatsApp users could only receive messages via Afrikaans, English and isiZulu. Regardless of the language men chose to receive messages in, clients could send messages in any language at any time with any text-based content.

For clinic staff and clinicians, 2wT requires smartphones, tablets, or desktop computers, all provided by the study team. A hub-and-spoke model was utilized during this intensive expansion phase with sites or general practitioners (GPs) acting as spokes and a study-funded nurse performing the role of the 2wT hub. Spokes were routine VMMC service delivery locations with non-study clinical and support teams who implemented VMMC in alignment with National Department of Health (NDoH) standards. Spokes completed 2wT duties including recruitment, enrollment, post-operative 2wT education, and 2wT client in-person follow-up as needed and completing referrals to care in the 2wT system. A study-specific VMMC nurse served as the central 2wT hub, providing text-based support with all 2wT participants enrolled at any site for wound care. The hub nurse triaged clients via messaging to in-person care, referring them to sites when needed. The hub nurse typically responded during routine weekday hours and, during campaign periods, on weekends. Embedded 2wT decision-support prompts for the hub nurse alerted to clients who did not respond by day 3 (minors) or adults (day 8) to reduce LTFU. The hub completed initial phone call-based tracing of 2wT participants who had not responded, referring them to sites for in-person tracing if not found. The hub nurse did not interact with, nor provide care for, SoC clients.

### Study design

The expansion study was implemented in two phases. The first, the subject of this analysis, was a modified, practical, SWD to expand 2wT access over time and setting, adapting the typical cluster randomized design to accommodate practical issues in determining the order of facilities and teams in routine VMMC service delivery. A mix of routine VMMC locations, including urban clinics, rural mobile team catchment areas, and general practitioner clinics (GPs) were targets for a diversity of implementation sites. To accommodate the pace of VMMC scale-up expected by the national VMMC donor, a compressed SWD was planned: Step 1 was intended to begin offering 2wT in three urban sites in one district on January 1; Step 2 was to expand to general practitioners (GPs) in a second district who would offer 2wT to clients starting on March 1; Step 3 was to add rural catchment areas from a mobile VMMC team in a third district starting to offer 2wT on May 1; and all sites would include an option of 2wT messaging platform (Short Message Service [SMS] or WhatsApp) to all 2wT participants starting on July 1 as Step 4. Men who enrolled at facilities before the intervention was available or who elected not to opt-into 2wT, received routine, SoC follow-up with in-person reviews scheduled in accordance with NDoH and local VMMC implementing partner requirements on post-operative days 2, 7, and 14 after VMMC.

During the SWD, a study-supported 2wT team provided quality assurance and clinical oversight, including the 2wT hub functions, considered an “intensive phase” of expansion. After completion of the SWD, a “maintenance phase” was initiated without dedicated 2wT study team support on January 1, 2024. The maintenance phase will continue through December 31, 2024, at which time the system and approach will transition to local teams, independently. Using the RCT as our guide, and recognizing complexity in routine, versus research settings, we hypothesized the 2wT would reach 60% of eligible clients and double the AE ascertainment rate over SoC.

### Implementation science framework

For the SWD study, we use the RE-AIM framework to analyze findings and understand facilitators and barriers to 2wT in South Africa (20, 21). For this quantitative component, we assess reach as the proportion of eligible clients who opted into the 2wT intervention, aiming to understand where we surpassed or failed to reach expected enrollment targets. We examined effectiveness using two common quality of VMMC care markers(4): safety and efficiency. We compared safety, as defined by the AE ascertainment rate, and efficiency, as the number of in-person follow-up visits, between intervention and comparison SoC clients. Examination of these two constructs inform potential 2wT expansion and aim to provide evidence for 2wT as an approach to augment SoC follow-up for interested and eligible men. Adoption, implementation and maintenance aspects of the framework are explored in a parallel qualitative manuscript (22).

### SWD implementation

The study started on January 1^st^, 2023 and enrollment concluded on October 19^th^, 2023. Clients were followed through December 2023 in case late AEs were reported. Local VMMC site teams were trained on the use of 2wT by the study team: A one-day training was conducted for each site and each team in a mix of theory and practice-based activities and included follow-up mentoring via WhatsApp and site visits over the course of the study. The training was repeated as refresher training as needed to reduce the impact of VMMC mobility and job transfers.

### Interventions: 2wT and standard of care (SoC)

#### 2wT opt-in clients (intervention)

Males ages 15 and older who opted into 2wT received an enrollment message in front of the enrollment nurse to confirm 2wT participation and were counseled about the 2wT intervention, including how to respond to message prompts on their healing, before leaving the VMMC site. Confirmation of the functionality of the supplied phone number was done immediately through sending a welcome message to the enrolled participant. If a 2wT client responded with a potential complication, a VMMC nurse exchanged texts through the 2wT platform with the client to determine the symptoms, frequency, and severity, referring to care if needed. If 2wT participants did not respond to any text by day 3 for minors or day 8 for adults, clients were traced first by text, then phone, and then referred to in-person tracing as per NDoH guidelines (health care workers went to look for men at their homes). 2wT is free for clients to send and receive messages via SMS; WhatsApp requires client-paid data to work.

#### Client standard of care (SoC) VMMC follow-up (comparison)

Males ages 15 and older in the comparison group and non-intervention periods were scheduled for routine, in-person follow-up on post-surgery days 2, 7 and 14 (Table 1) (23). Clients who did not return to the clinic for follow-up on Day 2 or Day 7 were referred for tracing by phone or home visit according to NDoH guidelines.

**Table 1.** 2wT and SoC VMMC procedures overview.

| Specific Activities | 2wT | Routine |
| --- | --- | --- |
| Day 0 (Day of VMMC) |  |  |
| Routine VMMC surgery and counseling | ✓ | ✓ |
| VMMC procedure consent | ✓ | ✓ |
| Wound care/bandage removal counseling | ✓ | ✓ |
| Post-operative 2wT education | ✓ |  |
| Scheduled in-person follow-up |  |  |
| Routine Day-2 |  | ✓ |
| Routine Day-7 |  | ✓ |
| Routine Day-14 |  | ✓ |
| Routine tracing if no 2wT or in-person visit | ✓ | ✓ |
| 2wT-specific processes |  |  |
| Response requested prompts: post-operative days 1-3, 5, 7, 10 and 13 | ✓ |  |
| Educational messages on VMMC wound care: post operative days 4, 6, 8, 9, 11 and 12 | ✓ |  |
| NDoH routine procedures |  |  |
| In-person, any day, review for AE concern | ✓ | ✓ |
| Emergency VMMC after-hours care | ✓ | ✓ |
| AE type, severity, and management documented | ✓ | ✓ |
| All VMMC reporting on routine NDoH forms | ✓ | ✓ |

#### All clients

All clients received routine pre-operative counseling, HIV testing, surgical VMMC, post-operative counseling on bandage removal, wound care, and an emergency number to call for concerns, all in line with NDoH protocols (24). Clients in either follow-up method could attend visits for wound care support or suspicion of AEs at any healthcare facility at any time. AE identification, management, and reporting adhered to NDoH standard of care (25).

### Recruitment, eligibility, and enrollment

The SWD study aimed to enroll at least 800 men who opted into 2wT, with the goal of 200 participants per step. For 2wT participants, eligibility included: 1) ≥15 years; 2) possession of own phone at enrollment; 3) willing to respond to daily text; 4) provide contact details (phone, address); 5) completes surgical VMMC; 6) willing to follow NDoH VMMC protocols; 7) no interoperative AE; 8) receives confirmed 2wT enrollment text 9) literacy and digital literacy. Each site operationalized demand creation, eligibility verification, education, and enrollment procedures aligned with their specific setting and population. Enrollment was undertaken before or after VMMC as per routine site flow using either a phone/tablet app or the laptop-based system. Those who enrolled before VMMC, but had an intraoperative AE, were reverted to SoC in-person reviews.

### Power and sample size

The sample size calculation was based on comparing SoC care to 2wT using the AE rates from the randomized control trial (9). The sample size calculation assumed that with 200 men per zone, per quarter, if SoC care AE ascertainment = 0.5%, and 2wT= 2.0%, depending on how correlated AE rates are within zones, we expect power between 81.8% and 84.9% to establish that AE ascertainment is superior using 2wT. There was no upper bound on enrollment as 2wT uptake is influenced by routine care processes.

**Table 2.**
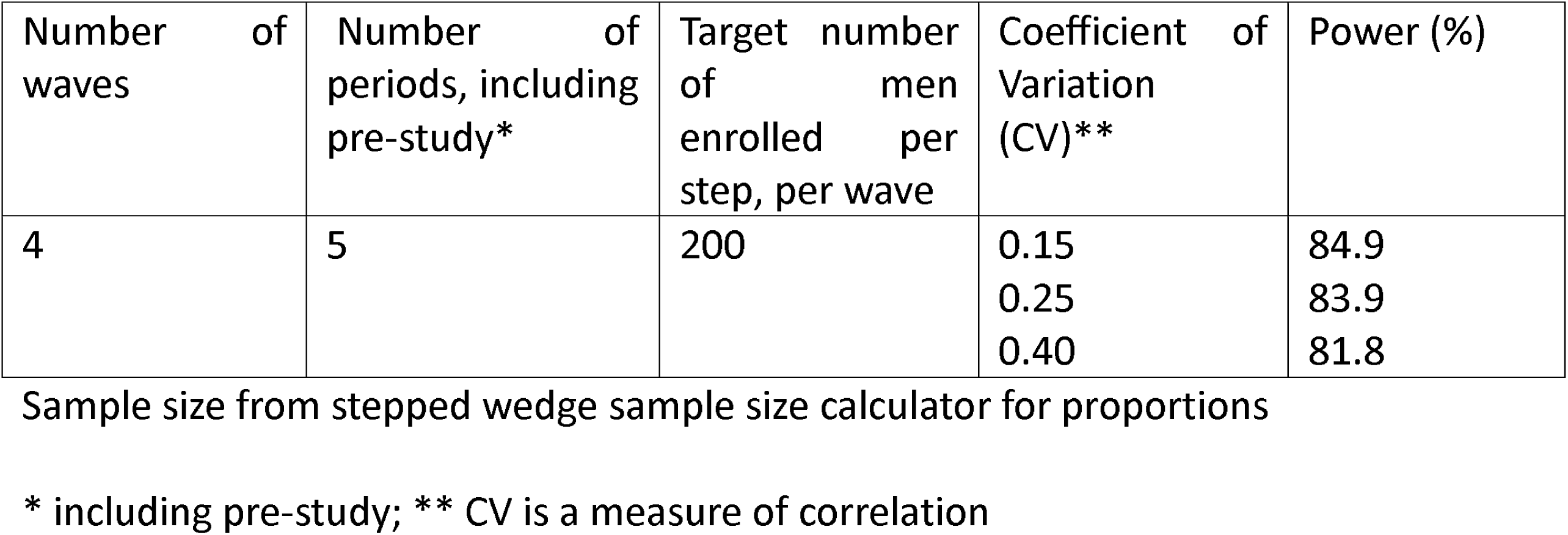
Power and sample size calculations.

| Number of waves | Number of periods, including pre-study* | Target number of men enrolled per step, per wave | Coefficient of Variation (CV)** | Power (%) |
| --- | --- | --- | --- | --- |
| 4 | 5 | 200 | 0.15<br>0.25<br>0.40 | 84.9<br>83.9<br>81.8 |

Sample size from stepped wedge sample size calculator for proportions
\* including pre-study; \*\* CV is a measure of correlation

### Outcomes

The primary outcomes of interest were reach and effectiveness (safety and workload) were maintained from the randomized controlled trial (RCT) (9). Reach is defined as: the proportion of VMMC clients who opted into 2wT/all clients, by step and overall. The primary safety outcome was the cumulative AE ascertainment rate, defined as the # of combined moderate or severe AEs/ total number of males who had any follow-up within 14 days of VMMC (clinic visit or 2wT response, by arm). AE *ascertainment* includes: confirming, managing, following up on, and documenting AEs. The workload outcome was defined as: the average # of in-person follow-up visits compared between SoC and 2wT-based follow-up. Severe AEs are those that require surgical intervention or hospitalization due to surgical or post-operative healing complications. Any AE not classified as severe, but which requires intervention by a health care provider or medication, is considered moderate(25). Secondary outcomes included: timing and severity of AEs; 2wT response rates; response type (potential AE, no AE, no response or free text (a non-numeric response); uptake by clients and site; tracing; and LTFU.

### Data sources

Data abstracted from routine VMMC forms for 2wT participants included: visit dates, AE type, AE severity, and clinical notes on healing. This data was captured and stored in a secure data management system and extracted in de-identified form for analysis. Supplemental 2wT database data included: daily text response, potential self-reported AEs, client messages exchanged with Hub nurse, Hub referrals for clients to site for review, and sites tracing clients who did not respond via SMS. For workload, visit attendance data was obtained from 2wT database and NDoH forms.

### Data Analysis

A descriptive analysis was performed to examine differences in patient, intervention, implementation, and safety and efficiency variables between the baseline and intervention periods, and between 2wT and SoC clients during intervention periods. We used multivariate generalized linear mixed-effects models (GLMM) to examine the odds ratio (OR) of having no visits and the mean number of in-person visits between intervention period and baseline period; between 2wT and SoC during the intervention period; and between people who used WhatsApp and SMS among 2wT participants. We modeled clustering across the 6 sites using a site-level random effect and adjusted for confounding between intervention and time by including calendar year quarter as a fixed effect in the GLMMs. The AE ascertainment rates were estimated and reported with the associated 95% confidence intervals (CIs) assuming a binomial distribution within each subgroup. Fisher’s exact tests were used to test for statistically significant differences in AE ascertainment rates between subgroups. All analyses were performed using R version 4.3.2 with α = 0.05 as the level of statistical significance.

### Ethics

The internal review boards (IRB) of the University of Washington (STUDY00009703; CF) and the University of Witwatersrand, Human Research Ethics Committee (No. 200204; GS) approved the study protocol for the routine scale up of 2wT as part of the stepped wedge study. The VMMC procedure has a separate consent from the 2wT study. As 2wT during this stepped wedge expansion phase was implemented as part of routine VMMC follow-up, IRB approval was obtained to waive written and oral consenting for 2wT participants ages 15 and older to opt into the 2wT follow-up approach. No client IDs were included in the analyzed data.

## Results

### SWD Implementation

The SWD study started January 1, 2023. Step 1 started on January 30, launching 2wT at two urban sites in District 1. Step 2 started on April 24 in another urban site in District 1. On May 16, Step 3 began with two general practitioners (GPs) in District 2 and one rural mobile VMMC team in District 3. On August 17, Step 4 was initiated signaling that all new 2wT enrollees at any existing intervention location from Steps 1-3 were offered a choice of 2wT platform: SMS or WhatsApp.

### Primary Outcomes

#### Reach: 2wT enrollment

A total of 8444 men aged 15 years and older were circumcised during the study period. Of these, 1602 (19%) were in the baseline period (pre-study SoC) and 6842 (82%) were in the intervention period. Of the 6842 VMMCs over implementation waves, 2586 (37.8%) opted into 2wT (Table 3).

**Table 3.** Study wave start dates and enrollment.

| SWD Wave location | Start date | Setting | Baseline SoC period | Intervention period enrollment |  |  |  |  |
| --- | --- | --- | --- | --- | --- | --- | --- | --- |
|  |  |  |  | 2wT platform |  |  | SoC | Total |
|  |  |  |  | SMS | Whats App* | Total 2wT |  |  |
| Step 1: | Jan 30 | 2 urban | 150 | 1657 | 167 | 1824 | 982 | 2956 |
| Step 2: | Apr 24 | 1 urban | 247 | 152 | 0 | 152 | 529 | 928 |
| Step 3: | May 16 | 2 GPs | 0 | 149 | 0 | 149 | 482 | 631 |
|  |  | 1 rural | 1205 | 449 | 12 | 461 | 2263 | 3929 |
| TOTAL |  |  | 1602 | 2407 | 179 | 2586 | 4256 | 8444 |

\*WhatsApp as Step 4 was offered in addition to SMS to all enrolling 2wT participants across all sites starting August 17<sup>th</sup>, 2023.
SWD: Stepped wedge design; 2wT: two-way texting; SoC: Standard of Care; GPs: General Practitioners

##### Demographics

During the SWD, the majority of SoC participants were aged 30 or older (48.6%); 2wT participants were younger with the majority aged 15-29 years (66.2%). Most 2wT participants (70.5%) originated from urban facilities; whilst the majority (76.9%) of SoC participants came from non-urban clinics. English was the primary language selected (74%) among those who chose 2wT. As expected, most 2wT participants had no visits (94.2%) while most SoC clients (67.4%) reported 2 visits. On average, 2wT participants had a mean of 0.07 visits (SD 0.30) compared to SoC clients with a mean of 2.04 visits (SD 0.75). Over 99% of clients did not have a reported AE. Of 37 reported AEs during the study, 28 (80%) were reported among 2wT participants.

**Table 4:**
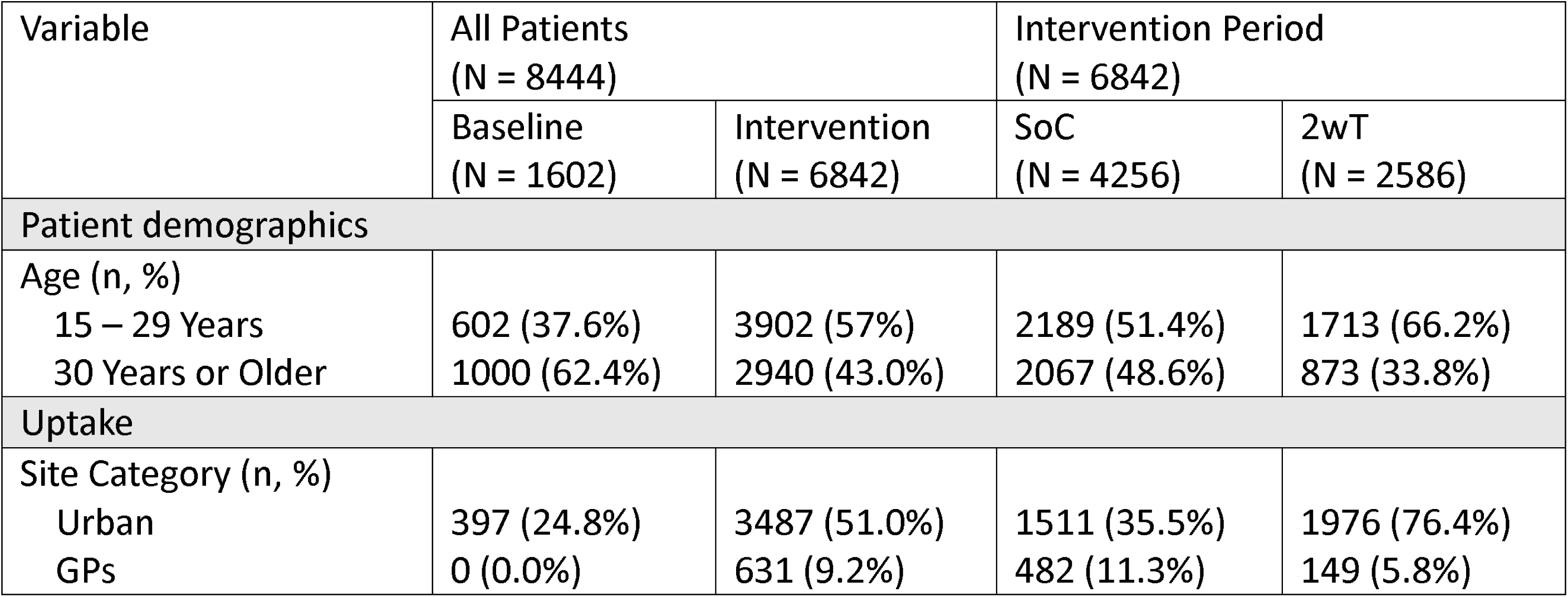
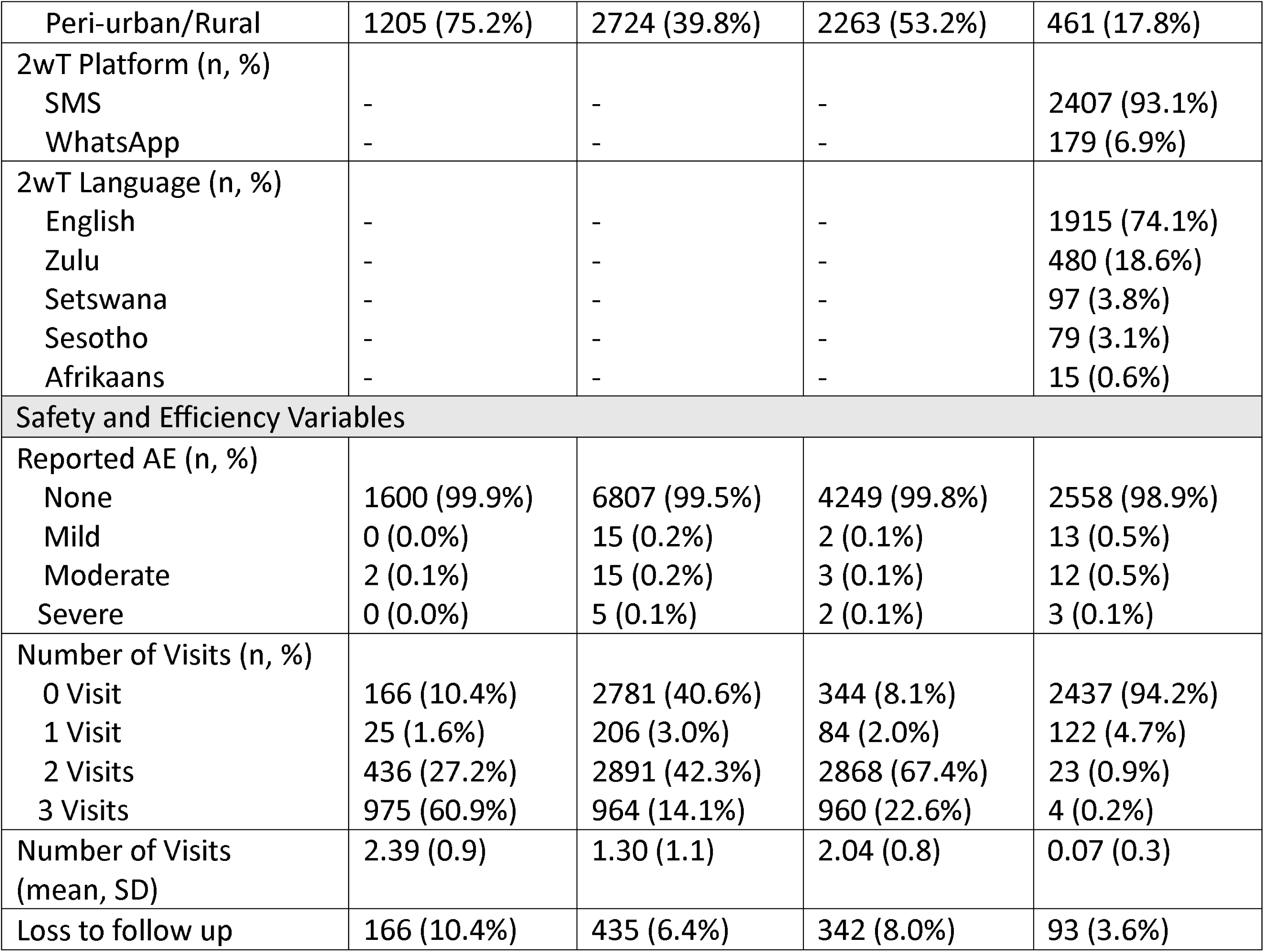
Descriptive statistics of patients, implementation, safety and efficiency.

| Variable | All Patients<br>(N = 8444) |  | Intervention Period<br>(N = 6842) |  |
| --- | --- | --- | --- | --- |
|  | Baseline<br>(N = 1602) | Intervention<br>(N = 6842) | SoC<br>(N = 4256) | 2wT<br>(N = 2586) |
| Patient demographics |  |  |  |  |
| Age (n, %) |  |  |  |  |
| 15 – 29 Years | 602 (37.6%) | 3902 (57%) | 2189 (51.4%) | 1713 (66.2%) |
| 30 Years or Older | 1000 (62.4%) | 2940 (43.0%) | 2067 (48.6%) | 873 (33.8%) |
| Uptake |  |  |  |  |
| Site Category (n, %) |  |  |  |  |
| Urban | 397 (24.8%) | 3487 (51.0%) | 1511 (35.5%) | 1976 (76.4%) |
| GPs | 0 (0.0%) | 631 (9.2%) | 482 (11.3%) | 149 (5.8%) |
| Peri-urban/Rural | 1205 (75.2%) | 2724 (39.8%) | 2263 (53.2%) | 461 (17.8%) |
| 2wT Platform (n, %) |  |  |  |  |
| SMS | - | - | - | 2407 (93.1%) |
| WhatsApp | - | - | - | 179 (6.9%) |
| 2wT Language (n, %) |  |  |  |  |
| English | - | - | - | 1915 (74.1%) |
| Zulu | - | - | - | 480 (18.6%) |
| Setswana | - | - | - | 97 (3.8%) |
| Sesotho | - | - | - | 79 (3.1%) |
| Afrikaans | - | - | - | 15 (0.6%) |
| **Safety and Efficiency Variables** |  |  |  |  |
| Reported AE (n, %) |  |  |  |  |
| None | 1600 (99.9%) | 6807 (99.5%) | 4249 (99.8%) | 2558 (98.9%) |
| Mild | 0 (0.0%) | 15 (0.2%) | 2 (0.1%) | 13 (0.5%) |
| Moderate | 2 (0.1%) | 15 (0.2%) | 3 (0.1%) | 12 (0.5%) |
| Severe | 0 (0.0%) | 5 (0.1%) | 2 (0.1%) | 3 (0.1%) |
| Number of Visits (n, %) |  |  |  |  |
| 0 Visit | 166 (10.4%) | 2781 (40.6%) | 344 (8.1%) | 2437 (94.2%) |
| 1 Visit | 25 (1.6%) | 206 (3.0%) | 84 (2.0%) | 122 (4.7%) |
| 2 Visits | 436 (27.2%) | 2891 (42.3%) | 2868 (67.4%) | 23 (0.9%) |
| 3 Visits | 975 (60.9%) | 964 (14.1%) | 960 (22.6%) | 4 (0.2%) |
| Number of Visits (mean, SD) | 2.39 (0.9) | 1.30 (1.1) | 2.04 (0.8) | 0.07 (0.3) |
| Loss to follow up | 166 (10.4%) | 435 (6.4%) | 342 (8.0%) | 93 (3.6%) |

#### Effectiveness

##### Safety

Only males who had any reported follow-up within 14 days were included in the safety analysis. Among the 2586 2wT participants, 93 (3.6%) did not respond to any messages and were not traced successfully or confirmed to have an AE, classified as LTFU. Among the 4256 SoC clients during the intervention period, 342 (8.0%) were classified similarly as LTFU. Therefore, 2493/2586 2wT (96.4%) and 3914/4256 SoC (92.0%) clients were included in safety analysis. Among those included clients with any post-operative follow-up after VMMC, the moderate or severe AE ascertainment rate was significantly greater among 2wT than SoC participants (0.60% vs. 0.13%, p = 0.0018) (Table 3).

**Table 5.**
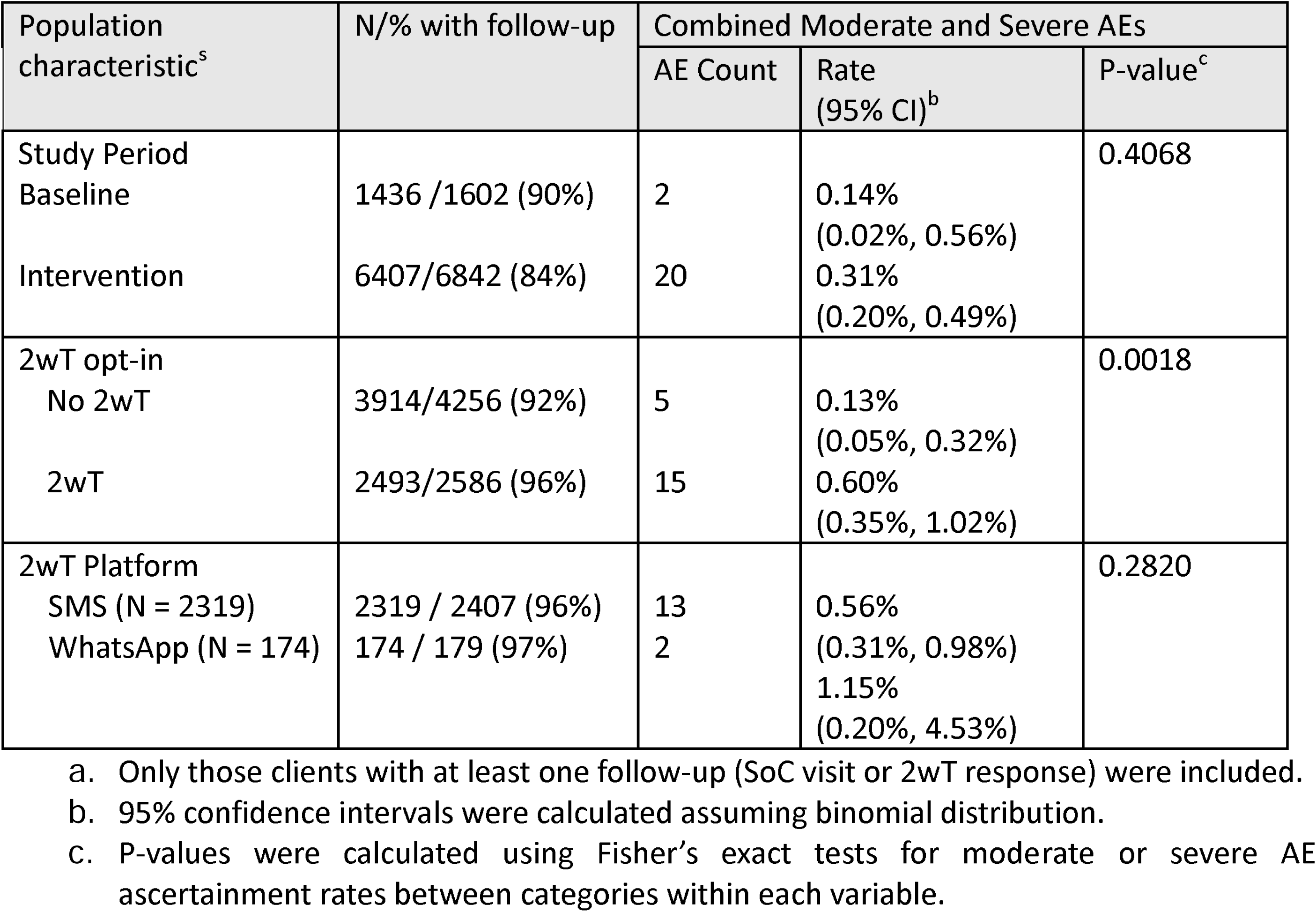
AE ascertainment rate by arm.

##### Efficiency

During the intervention period, 2wT participants were 12 times more likely to have zero visits as compared to SoC clients. In other words: 2wT participants had 2 fewer visits (mean = 2.09) than SoC clients.

**Table 6.** 2wT efficiency gains: probability for no-visit and mean visits by arm.

| Variable | Probability for No-visit<sup>a</sup> |  |  | Difference in mean visits<sup>b</sup> |  |  |
| --- | --- | --- | --- | --- | --- | --- |
|  | All clients<br>N=8444 | Intervent.<br>period<br>N=6842 | 2wT<br>N = 2586 | All clients<br>N=8444 | Intervent.<br>period<br>N = 6842 | 2wT<br>N = 2586 |
| Intervention vs. baseline | 2.97***<br>(2.5, 3.6) | - | - | -0.93***<br>(-1.0,-0.9) | - | - |
| Opted-in 2wT vs. No 2wT | - | 11.98***<br>(10.6,13.5) | - | - | -2.09***<br>(-2.1,-2.1) | - |
| WhatsApp vs. SMS | - | - | 1.00<br>(0.8, 1.2) | - | - | 0.03<br>(-0.0, 0.1) |

\* p<0.05 \*\*p<0.01 \*\*\*p<0.001
a. Estimations were based on multivariate generalized linear mixed-effects models with a logit link that included quarter as a fixed effect and site as a random effect.
b. Estimations were based on multivariate linear mixed-effects models that included quarter as a fixed effect and site as a random effect.

### Secondary outcomes

#### Timing and severity of AEs

During the intervention period, there were 15 moderate and severe AEs among 2wT participants: 12 moderate (3 bleeding; 3 infections; 6 wound disruption) and 3 severe AEs (2 bleeding and 1 wound disruption) (Table 7). Among SoC participants, there were 5 moderate (3 bleeding, 2 wound disruption) and 2 severe AEs (1 swelling, 1 wound infection). Only three males age ≤18 were identified with an AE, all were moderate AE among SoC males. For AEs identified on or before day 7, 4/7 AEs (57%) were identified among 2wT males and 3/7 AEs (43%) were identified among SoC men. Post-operative AEs beyond 14 days after circumcision procedure were identified among 4 males, all in 2wT. The median day for which the AE was detected for the 2wT group was 11 and for the SoC group was 9. Among those 2wT participants with an AE, 12/15 (80%) responded at least once in 14 days and 9/12 (75%) of them had reported a potential AE via the 2wT system. All AEs were related to VMMC and all occurred on or before day 37 post-operatively. All AEs were successfully resolved.

**Table 7:**
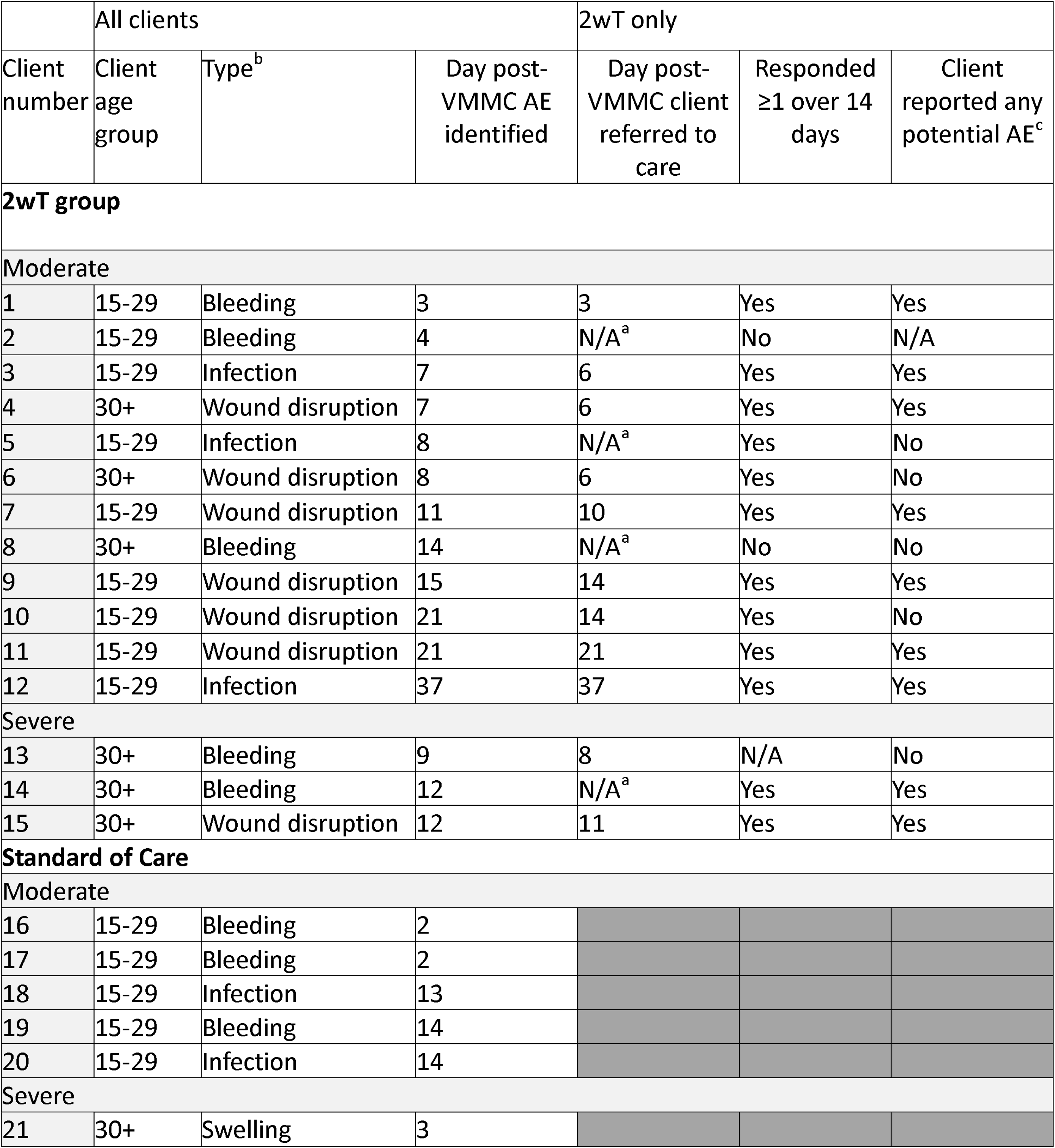
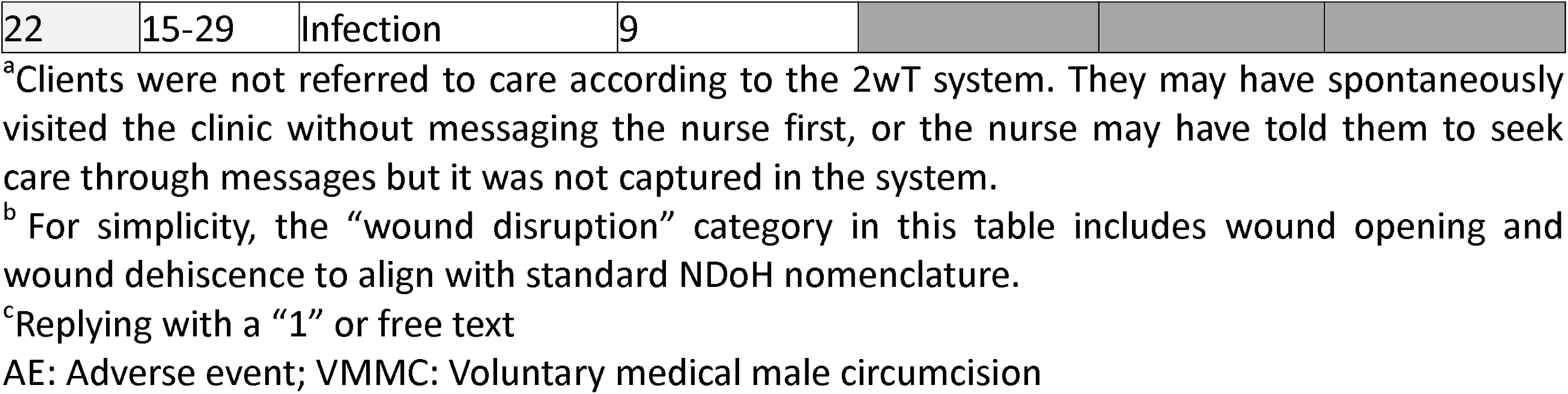
Description of study AEs by arm.

|  | All clients |  |  | 2wT only |  |  |
| --- | --- | --- | --- | --- | --- | --- |
| Client number | Client age group | Type<sup>b</sup> | Day post-VMMC AE identified | Day post-VMMC client referred to care | Responded ≥1 over 14 days | Client reported any potential AE<sup>c</sup> |
| **2wT group** |  |  |  |  |  |  |
| Moderate |  |  |  |  |  |  |
| 1 | 15-29 | Bleeding | 3 | 3 | Yes | Yes |
| 2 | 15-29 | Bleeding | 4 | N/A<sup>a</sup> | No | N/A |
| 3 | 15-29 | Infection | 7 | 6 | Yes | Yes |
| 4 | 30+ | Wound disruption | 7 | 6 | Yes | Yes |
| 5 | 15-29 | Infection | 8 | N/A<sup>a</sup> | Yes | No |
| 6 | 30+ | Wound disruption | 8 | 6 | Yes | No |
| 7 | 15-29 | Wound disruption | 11 | 10 | Yes | Yes |
| 8 | 30+ | Bleeding | 14 | N/A<sup>a</sup> | No | No |
| 9 | 15-29 | Wound disruption | 15 | 14 | Yes | Yes |
| 10 | 15-29 | Wound disruption | 21 | 14 | Yes | No |
| 11 | 15-29 | Wound disruption | 21 | 21 | Yes | Yes |
| 12 | 15-29 | Infection | 37 | 37 | Yes | Yes |
| Severe |  |  |  |  |  |  |
| 13 | 30+ | Bleeding | 9 | 8 | N/A | No |
| 14 | 30+ | Bleeding | 12 | N/A<sup>a</sup> | Yes | Yes |
| 15 | 30+ | Wound disruption | 12 | 11 | Yes | Yes |
| **Standard of Care** |  |  |  |  |  |  |
| Moderate |  |  |  |  |  |  |
| 16 | 15-29 | Bleeding | 2 |  |  |  |
| 17 | 15-29 | Bleeding | 2 |  |  |  |
| 18 | 15-29 | Infection | 13 |  |  |  |
| 19 | 15-29 | Bleeding | 14 |  |  |  |
| 20 | 15-29 | Infection | 14 |  |  |  |
| Severe |  |  |  |  |  |  |
| 21 | 30+ | Swelling | 3 |  |  |  |
| 22 | 15-29 | Infection | 9 |  |  |  |

<sup>a</sup>Clients were not referred to care according to the 2wT system. They may have spontaneously visited the clinic without messaging the nurse first, or the nurse may have told them to seek care through messages but it was not captured in the system.
<sup>b</sup> For simplicity, the “wound disruption” category in this table includes wound opening and wound dehiscence to align with standard NDoH nomenclature.
<sup>c</sup>Replying with a “1” or free text
AE: Adverse event; VMMC: Voluntary medical male circumcision

#### Response rates and clinic / provider uptake

Among those opting into the 2wT approach, 2wT client response rates varied by sites. Response rates by day 3 and by day 14 were generally high among 2wT men, ranging from 40%-86% on day 3 and 60%-90% by day 14. Most males responded in the morning soon after receipt of the response-requested message. Considering the location of 2wT client enrollment, 2wT opt-in proportions varied greatly by enrollment location, ranging from a high of 68% 2wT enrollment at site 1, an urban clinic, and a low of 17% at site 6, in a mobile VMMC team in a rural catchment area.

**Table 8:** 2wT uptake: 2wT participant response rates and 2wT adoption rates.

| Step * | Sites | 2wT client response rates |  | 2wT adoption rates during intervention |  | Total clients per site during intervention |
| --- | --- | --- | --- | --- | --- | --- |
|  |  | By Day 3 | By Day 14 | 2wT N/% | SoC N/% |  |
| Step 1 urban | 1 | 738 (73%) | 819 (81%) | 1014 (68%) | 474 (32%) | 1488 |
|  | 2 | 602 (74%) | 680 (84%) | 810 (61%) | 508 (39%) | 1318 |
| Step 2 urban | 3 | 131 (86%) | 137 (90%) | 152 (22%) | 529 (78%) | 681 |
| Step 3 GPs / rural | 4 GP | 33 (66%) | 39 (78%) | 50 (20%) | 196 (80%) | 246 |
|  | 5 GP | 45 (46%) | 59 (60%) | 99 (26%) | 286 (74%) | 385 |
|  | 6 | 283 (61%) | 335 (73%) | 461 (17%) | 2263 (83%) | 2724 |
|  | Total | 1832 (71%) | 2069 (80%) | 2586 (38%) | 4256 (62%) | 6842 |

\*WhatsApp as Step 4 was offered in addition to SMS to all enrolling 2wT participants across all sites starting August 17<sup>th</sup>, 2023. These participants are included in their location of enrollment.

##### 2wT response types

On days that a response was requested, an average of 45% responded. On any given day, most 2wT participants did not respond. Response rates declined over the 14 days. However, of 2586 2wT males, 80% responded at least once over the follow-up 14 days and 863 (33%) sent at least one potential AE response to interact with the nurse and receive support. Only 517 2wT participants (20%) never responded over 14 days; 76% were traced successfully and without an AE.

**Figure 1:**
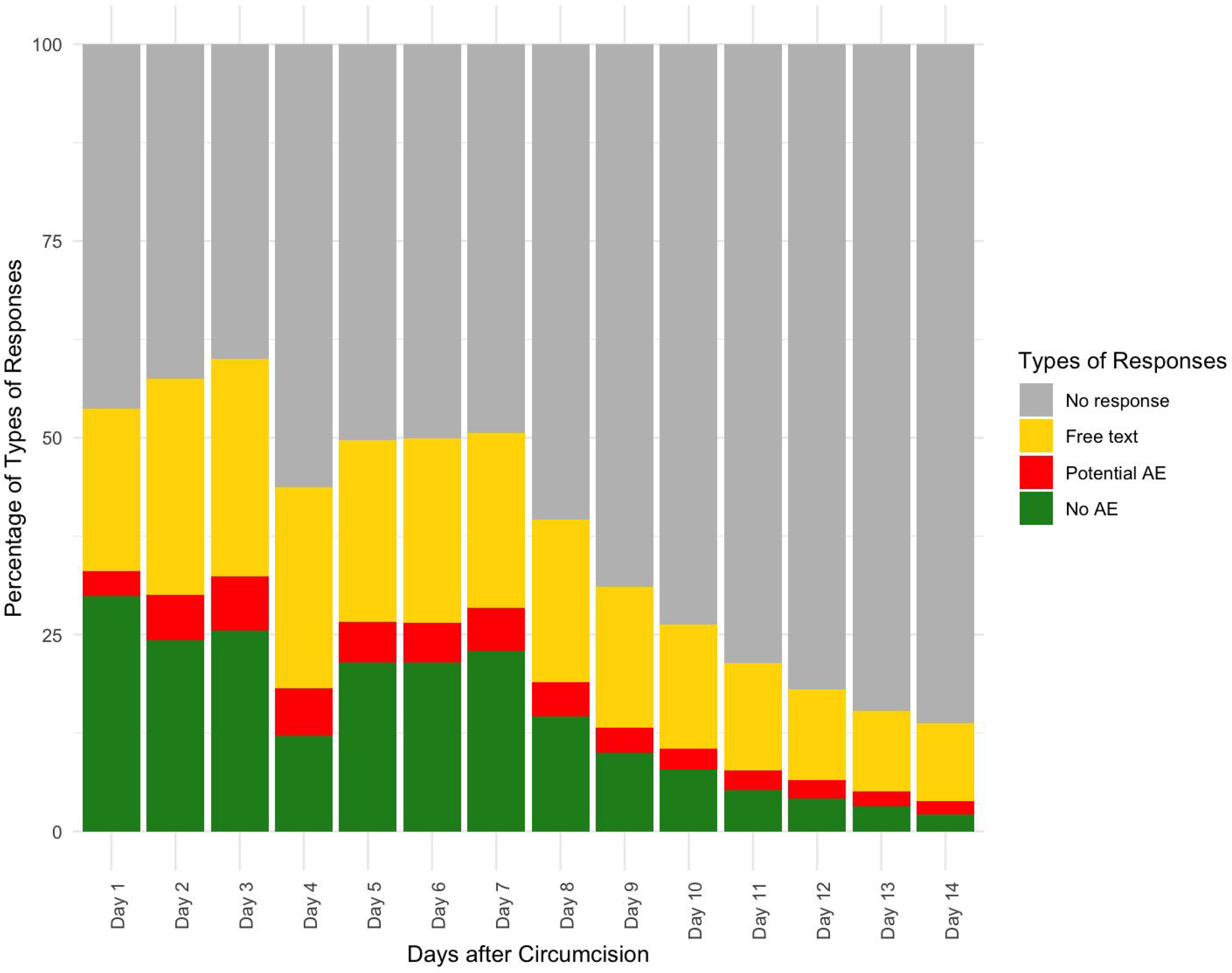
2wT response types over 14 follow-up days

## Discussion

The study provides evidence that 2wT for VMMC follow-up in South Africa reaches a sizeable proportion of VMMC clients and is an effective approach for VMMC follow-up, reducing visits and increasing AE ascertainment. Participants using 2wT are engaged in their care and intentional in seeking 2wT-based support or following up on referrals to care. The 2wT follow-up approach ascertained nearly four times as many AEs as the routine, in-person visit approach, confirming our hypothesis and suggesting clear advantages for client safety using 2wT. While demonstrating quality care advantages in identifying, referring, and documenting AEs, the 2wT approach also reduced in-person visits dramatically, encouraging only those few with a desire or need to return for in-person reviews. Response rates among 2wT participants for days a response was expected, averaged 45%, demonstrating engagement of 2wT participants with their caregivers. Despite these persuasive benefits to the 2wT follow-up approach, 2wT reach fell short of hypothesized reach of 60% likely due to lack of buy-in among sites and clinicians (22). We discuss opportunities, suggest possible constraints, and offer recommendations to create momentum for 2wT expansion.

First, similar to previous findings in Zimbabwe and South Africa (7, 9), 2wT is clearly effective to both AEs identification and reduction in unnecessary in-person reviews. This is an opportunity for more dissemination of these distinct advantages of the 2wT approach for follow-up. AEs are also more likely to be reported using 2wT as there is extensive documentation of client-to-clinician interaction, referrals to care, and visits. While AEs could be undercounted in either group as clients may seek care in any location and AEs may be treated but unreported, this is still persuasive evidence that 2wT improves AE ascertainment and, therefore, reporting. Interestingly, there were more AEs identified after day 14 among 2wT participants compared to SoC participants. While these participants could have delayed care seeking, that is unlikely as each of these four clients responded via 2wT within 14 days and most were referred to care by the 2wT nurse. To improve AE reporting, an automated message could be sent to 2wT participants referred to care requesting a response on whether they had a visit, triggering the 2wT hub to solicit additional details. This innovation could further identify potentially missed AEs, providing data for AE triangulation and data quality. For visit efficiency, it is also clear that 2wT participants required or desired few in-person reviews. While over or undercounting visits is also possible in either arm, the 2wT efficiency gains from fewer in-person reviews are undeniable.

Second, although clients who opt into 2wT are highly responsive and engaged in the approach, 2wT is not achieving expected reach: only 38% of participants during the intervention period chose to enroll. This is a clear constraint to expansion. Previously, when 2wT recruitment was implemented by the RCT study team, and not routine VMMC service delivery teams, reach was 75% (9). Among those reached in the Zimbabwe or South Africa RCT and in the SWD, clients responded well and were engaged in their follow-up, showing that clients largely approve of the approach. Lower than anticipated 2wT reach may be influenced by client-side or clinic-side constraints. VMMC clients may lack awareness of the 2wT approach, lack cell phones, not want to provide contact information, or not understand that participating in 2wT using SMS is free. To address these potential client-side constraints, we suggest increasing 2wT education, awareness, and promotion during routine VMMC demand creation activities, including use of improved educational posters and short promotional videos. Further, clinic- and clinician-side barriers may curtail expansion momentum. Although the 2wT approach was optimized locally for VMMC healthcare workers using simple technology (14), routine VMMC teams may still consider 2wT to be additional effort. Not all site teams may offer 2wT to all eligible males or do so only during specific times. Not all VMMC follow-up visits may be completed as reported. Alternatively, even with clinic or clinician support, VMMC staff turnover and staff shortages can create bottlenecks in client flow or result in gaps in 2wT knowledge among clinic teams, potentially reducing 2wT integration into routine workflows. Innovations that streamline client enrollment via QR codes or allow clients to enroll later from home could reduce the enrollment burden, improving 2wT enthusiasm. Broader dissemination of efficiency gains due to reduced unnecessary visits may also motivate clinic teams to adopt 2wT. Additional refresher training and follow-up mentoring by 2wT champions could also help encourage site teams to become comfortable and confident in the 2wT approach.

Lessons from another mobile health intervention may offer suggestions to improve 2wT expansion. MomConnect, a digital health initiative launched by the South African Department of Health in 2014 aims to provide education to pregnant women and new mothers (26). Although MomConnect lacks rigorous evidence of impact, the intervention scaled nationally to reach millions of South African women (27). MomConnect and 2wT share several crucial indicators of well-designed interventions, including: user-centered design to ensure health-related content was relevant, timely, and available in multiple languages; using SMS to include those with basic phones; include monitoring and evaluation processes for performance tracking; and supporting the local healthcare system via referrals and encouraging follow-up care in routine settings. Unlike 2wT, however, MomConnect has strong government support that enhances credibility and reach; widespread dissemination campaigns on radio and posters; and a robust funding base via public-private partnerships with mobile network operators, technology companies, non-governmental organizations (NGOs), and international donors. There may simply be less appetite for male-focused, HIV prevention interventions, less excitement about digital interventions focused on an acute or short engagement in care; or more momentum to foster ongoing reproductive education for adolescent females and women, factors that could curtail scale up potential (28).

To overcome these challenges and maintain the option of the 2wT approach within the routine VMMC services, several recommendations for 2wT scale-up should be considered. First, although 2wT appears cost-effective and technology costs are largely balanced by reductions in follow-up costs at scale, we note the need for sustained funding to support this augmentation of VMMC follow-up as part of large investments in quality service delivery. Second, with this preponderance of evidence, we advocate for a change in VMMC policy to accept telehealth as a complementary method of follow-up. However, even with revised President’s Emergency Plan for AIDS Relief (PEPFAR) guidance that supports virtual follow-up after VMMC for low risk clients (1), shrinking donor funds creates concern about guidelines that require in-person follow-up for all patients (4). This reluctance among program managers to defy current donor policies that require in-person reviews may stymie 2wT expansion enthusiasm. Third, although digital health interventions like 2wT may benefit the VMMC program, integration of 2wT into the current eHealth infrastructure requires formal approval from NDoH, international organizations, private sector stakeholders, and local implementing partners – an effort that is critical, but formidable. It is hoped that continued, robust 2wT monitoring and evaluation of 2wT will continue to provide valuable insights into what works and why, allowing for continuous improvement and adaptation of strategies that may lead to broader adoption at scale.

### Strengths and Limitations

The 2wT approach was implemented as opt-in, allowing for any client aged 15 and older with a phone (and who met eligibility guidelines) to participate within the routine VMMC program if desired. It is unknown whether those who did not opt-in were ineligible, or interested in 2wT. Ideally 2wT was offered to all men with phones over age 15 as part of routine follow-up – allowing men to opt-into 2wT instead of in-person visits. In practice, sites and teams enrolled males into 2wT when human resources or capacity were sufficient. The study was not implemented as expected due to changes in VMMC implementing partners and national VMMC priorities that altered the location, timing and order of steps, muddling evaluation of 2wT impact. Furthermore, the evaluation of WhatsApp as a 2wT delivery platform showed that men appreciated this option (29) but with the small sample size, we cannot compare outcomes by SMS or WhatsApp. AEs could be underreported in either group but are more likely to be missed in the routine arm. Similarly, visits could be missed in either arm as clients could seek follow-up care in any clinic. More likely, however, follow-up visit attendance rates may be overreported in the routine arm as seen in other VMMC programs operating at scale (30), creating a false comparison of follow-up visit workload. Lastly, the 2wT study team provided quality assurance throughout the study to ensure adherence to protocols, increasing supervision and support. This additional level of support requires additional human resources that should be considered for sustained, high-quality 2wT implementation.

## Conclusions

With now over five years of scale-up of the 2wT approach for VMMC follow-up from both South Africa and Zimbabwe, the evidence supports further adoption of 2wT for VMMC service delivery in routine settings. Noting that support from local ministries of health and government bodies are the first step towards expansion of the 2wT approach, VMMC guidelines and donor community reporting requirements should be updated in support of 2wT-based follow-up. These reforms would provide local authorities with reassurance that safety and efficiency can be improved or maintained with the option for telehealth follow-up alongside in-person visits. If approved, 2wT would provide a model for other services to improve male engagement in care (e.g., TB treatment, sexually transmitted infections) or other acute health concerns among other populations (childhood diseases, respiratory infections, post-operative care) that could similarly benefit from direct provider-to-client communication and improved care. With the existence of evidence for usability, cost-effectiveness, and safety, we call on implementing partners and collaborators to help create momentum for 2wT after VMMC and for use of this approach in other acute follow-up contexts.

## Supporting information

U Witwatersrand Ethical Approval

U Washington Ethical Approval

Data for routine clients

Data for 2wT clients

## Data Availability

All data produced in the present study are contained in the supplemental files uploaded to MedRxiv.

## Acknowledgements

The authors would like to thank the Departments of Health of the Gauteng and North West Provinces; implementing partner, Right to Care, for enabling participation of the clinical teams; men who participated in the expansion study, and the Aurum Institute study implementation team for their dedication and skill in study implementation. Thank you also to Femi Oni and Mourice Barasa Wafula for their work on the 2wT system and their dedication to ensuring it ran smoothly.

## Funding

Research reported in this publication was supported by the National Institute of Nursing Research (NINR) of the National Institutes of Health under award number 5R01NR019229, “Expanding and Scaling Two-way Texting to Reduce Unnecessary Follow-Up and Improve Adverse Event Identification Among Voluntary Medical Male Circumcision Clients in the Republic of South Africa.” Statistical support was provided by the University of Washington and its Fred Hutch Center for AIDS Research, a National Institutes of Health (NIH)-funded program under award number AI027757 that is supported by the following NIH Institutes and Centers: National Institute of Allergy and Infectious Diseases, National Cancer Institute, National Institute of Mental Health, National Institute on Drug Abuse, Eunice Kennedy Shriver National Institute of Child Health and Human Development, National Heart, Lung and Blood Institute, National Investigation Agency, National Institute of General Medical Sciences, and National Institute of Diabetes and Digestive and Kidney Diseases. The content is solely the responsibility of the authors and does not necessarily represent the official views of the National Institutes of Health.

## Conflicts of Interest

None declared.

## Notes

### Competing Interest Statement

The authors have declared no competing interest.

### Author Declarations

The Human Research Ethics Committee of the University of the Witwatersrand and the IRB of the University of Washington gave ethical approval for this work.

