## Supplementary material for "Strengthening evidence for text-based telehealth in post-operative care: A pragmatic study of the reach and effectiveness of two-way, text-based follow-up after voluntary medical male circumcision in South Africa": U Witwatersrand Ethical Approval

UNIVERSITY OF THE WITWATERSRAND, JOHANNESBURG

Human Research Ethics Committee: (Medical)
FWA Registered No IRB 00001223

**SECRETARIAT:** Suite 189, Private Bag x2600, Houghton 2041, South Africa Tel: +27-11-274 9200 Fax: +27-11-274 9281

29 June 2020 **EMAILED & COURIERED**

Prof KG Setswe,
The Aurum Institute
The Ridge, Aurum House
29 Queens Road
Parktown
2193

Dear Prof Setswe,

**PROTOCOL: AUR2-8-270 2WT - EXPANDING AND SCALING TWO-WAY TEXTING TO REDUCE UNNECESSARY FOLLOW-UP AND IMPROVE ADVERSE EVENT IDENTIFICATION AMONG VOLUNTARY MEDICAL MALE CIRCUMCISION CLIENTS IN REPUBLIC OF SOUTH AFRICA**

**ETHICS REFERENCE NO: 200204**

**RE : FINAL ETHICS APPROVAL**

This is to certify that the above-mentioned trial has been approved by the University of the Witwatersrand, Human Research Ethics Committee (HREC), and the Protocol/Expert Reviewer. Date of Meeting where trial was reviewed: 28 February 2020.

The University of the Witwatersrand, Human Research Ethics Committee Approval Granted for the above mentioned study is valid for five years. Where required by Sponsor to have approval on a more frequent basis it remains the responsibility of the Sponsor and Investigator to apply for continuing review and approval, or for the duration of the Trial.

**1. It is the responsibility of the Sponsor and Principal Investigator to ensure, where required, that relevant approvals are in place and compliance with the following is adhered to before a trial may begin:**

- If trial is being conducted in Provincial Health facilities: Approval from the Hospital CEO / Clinic Manager / District Research Committee (whichever is applicable) be obtained.
- The study is submitted onto The National Health Research Database (NHRD).
- The relevant approvals are uploaded onto the NHRD system: Ethics Approval, SAHPRA Approval, Hospital CEO / Clinic Manager / District Research Committee Approval.

* A copy of the SAHPRA Approval and/or SAHPRA Notification letter must be submitted to the Ethics Secretariat Office for record purposes (IF SAHPRA APPROVAL / NOTIFICATION IS APPLICABLE).

* The study is conducted according to the protocol submitted to the University of the Witwatersrand, Human Research Ethics Committee. Any amendments to the protocol must first be submitted to the Human Research Ethics Committee for approval.

* During the study, the University of the Witwatersrand, Human Research Ethics Committee is informed immediately of :

- Any Unexpected Serious Adverse Events or Unexpected Adverse Drug Reactions, which, in the Investigator and/or the Sponsor's opinion are suspected to be related to the study drug. (Refer to POL-IEC–001 and SOP-IEC–005, Item 3.4).
- Any data received during the trial which, may cast doubt on the validity of the continuation of the study.

* The University of the Witwatersrand, Human Research Ethics Committee is notified of any decision to discontinue the study and the reason stated.

* The Investigators authorised by this approval participate in this study. Additional Investigators shall be submitted to the University of the Witwatersrand, Human Research Ethics Committee for approval prior to their participation in the study.

* In the event of an authorised Investigator ceasing to participate in the study, the University of the Witwatersrand, Human Research Ethics Committee must be informed and the reason for such cessation given.

**2. PRINCIPLES OF INFORMED CONSENT:**

* The University of the Witwatersrand, Human Research Ethics Committee requires that in all studies, the Principles of Informed Consent are adhered to. This applies to volunteers as well as patients.

**3. PROGRESS REPORTS:**

* The University of the Witwatersrand, Human Research Ethics Committee requests that the SAHPRA Progress Reports be submitted twice a year either in March and September or six monthly from start of study to the HREC Secretariat Office - 011 274 9281 and a report of the final results, at the conclusion of the study. (IF APPLICABLE)

**4. REIMBURSEMENT TO PATIENTS FOR TRANSPORT:**

* The Human Research Ethics Committee: (Medical) is in agreement that reimbursement per visit is according to the South African Health Products Regulatory Authority and that reimbursement should be appropriate according to the situation.

**5. TRANSPORT AND STORAGE OF BLOOD AND TISSUE SAMPLES IN SOUTH AFRICA:**

* If blood specimens are to be stored for future analysis and is planned that such analysis will be done outside Wits, then the blood must be stored at a facility in South Africa agreed with the relevant IRB, with release of sub-samples only once projects have been approved by the local Research Ethics Committee applicable to where the analysis will be done as well as by the Wits Human Research Ethics Committee: (Medical).

**6. GENETIC TESTING:**

* The Human Research Ethics Committee: Medical; will not approve open-ended genetic testing as this does not fit the Human Research Ethics Committee criteria.

**7. GOOD CLINICAL PRACTICE:**

* The South African Department of Health, South African Health Products Regulatory Authority requires Good Clinical Practice (GCP) Training for all Investigators in Clinical Trials, and that GCP training be renewed every three (3) years.

As yet, there are no National Guidelines for the content of GCP courses. Until these are available the Wits Human Research Ethics Committee (Medical) will note courses completed by Investigators without approval of the content of the individual courses.

**8. THE SUPPORTING APPROVAL DOCUMENTS ARE ATTACHED:**

8.1 Ethics Approval Form signed by the Chairperson of the HREC - Kindly return the copy of the Approval Form signed by the Principal Investigator(s) per fax: 011 274 9281 for our records (this is applicable with the initial Approval).

8.2 List of members present at the HREC meeting held as per INDEPENDENT ETHICS COMMITTEE APPROVAL FORM

**9. WE AWAIT YOUR RESPONSES AS REQUESTED:** Ensure to have these documents forwarded at the earliest for the HREC records.

* SAHPRA Approval letter and/or letter of Notification before the above study may commence / or where an Amendment may be implemented (IF SAHPRA APPROVAL / NOTIFICATION IS APPLICABLE). It remains the responsibility of the Principal Investigator and/or Sponsor to ensure that the relevant approvals are in place.

* Copy of Independent Ethics Declaration Approval Form signed by the Principal Investigator. (this is applicable with the initial Approval).

* Kindly forward the above to the undersigned at fax: 011 274 9281 at your earliest convenience.

***Approved on the following conditions:***
***1. Please submit GCP and Ethics Training Certificates for Ms J Pienaar***

The above has been noted for the Ethics Committee information and records.

***KINDLY FORWARD TO THE RELEVANT INVESTIGATORS / CRA / SPONSOR / STUDY CO-ORDINATORS - WHERE APPLICABLE***

Regards,

**PROF CLEMENT PENNY**
For and on behalf of the Human Research Ethics Committee: (Medical)

UNIVERSITY OF THE WITWATERSRAND, JOHANNESBURG

### INDEPENDENT ETHICS COMMITTEE APPROVAL FORM

| | | | |
|---|---|---|---|
| **Ethics Reference No.** | **200204** | **Date of Meeting** | **28-Feb-2020** |
| | | **Recertification Due** | **26 February 2021 (If applicable)** |

| | | | |
|---|---|---|---|
| **Principal Investigators:** | Prof KG Setswe | **Investigators:** | Ms J Pienaar |
| **Co-Principal Investigator** | Prof CB Feldacker | | |

| | |
|---|---|
| **Protocol Title:** | Expanding and Scaling Two-way Texting to Reduce Unnecessary Follow-up and Improve Adverse Event Identification Among Voluntary Medical Male Circumcision Clients in Republic of South Africa |

| | **DOCUMENTS REVIEWED** | **Tick As Appropriate** | **Yes** | **No** |
|---|---|---|---|---|
| Protocol Number | **AUR2-8-270 2WT** | **Date:** 31-Jan-2020 | | |
| Protocol | Protocol AUR2-8-270 2WT, Version 1.0 | **Date:** 31-Jan-2020 | ☑ | ☐ |
| Investigator's Brochure | N/A - Version: - - Dated: | | ☑ | ☐ |
| Subject Information/Consent Form | Participant Enrolment Forms - Version: Version 1.0 - Dated: 31 Jan 2020 | | ☑ | ☐ |
| | Flow Chart - Version: Version 2.0 - Dated: 06 Mar 2020 | | ☑ | ☐ |
| | Information Sheet - Version: Version 2.0 - Dated: 06 Mar 2020 | | ☑ | ☐ |
| | Participant Recruitment and Information Sheet for Waterberg - Version: Version 2.0 - Dated: 06 Mar 2020 | | ☑ | ☐ |
| | Participant Information Sheet Form and Informed Consent Form for RCT - Version: Version 2.0 - Dated: 06 Mar 2020 | | ☑ | ☐ |
| | Participant Information Sheet and Informed Consent Form for Step-Wedge - Version: Version 2.0 - Dated: 06 Mar 2020 | | ☑ | ☐ |
| | Participant Information Sheet and Informed Consent Form for Pilot Study - Version: Version 2.0 - Dated: 06 Mar 2020 | | ☑ | ☐ |
| | Participant Information Sheet and Informed Consent Form for Healthcare Worker Interviews and Audiotaping - Version: Version 2.0 - Dated: 06 Mar 2020 | | ☑ | ☐ |
| Advertisements | | | | |
| Questionnaires | Interview Guide for Healthcare Workers - Version: Version 1.0 - Dated: 31 Jan 2020 | | ☑ | ☐ |
| | DSMB Charter - Version: Version 1.0 - Dated: 01 Nov 2018 | | ☑ | ☐ |
| | Recruitment script - Version: Version 1.0 - Dated: 31 Jan 2020 | | ☑ | ☐ |
| | Text language - revision/input requests - Version: Version 1.0 - Dated: 31 Jan 2020 | | ☑ | ☐ |
| | Usability survey for healthcare workers and pilot participants - Questions for Pilot Men - Version: Version 1.0 - Dated: 31 Jan 2020 | | ☑ | ☐ |
| | Standard operating procedures for texting and data - Version: Version 2.0 - Dated: 01 Jan 2020 | | ☑ | ☐ |
| | Standard operating procrdures for the clinical team - Version: Version 5.0 - Dated: 01 Jun 2018 | | ☑ | ☐ |
| | Usability survey for healthcare workers and pilot participants - Nurses/clinicians using the system to generate and respond to texts - Version: Version 1.0 - Dated: 31 Jan 2020 | | ☑ | ☐ |
| | VMMC text-based follow-up time and motion survey - Version: Version 1.0 - Dated: 31 Jan 2020 | | ☑ | ☐ |
| | Participant opinion survey - Version: Version 1.0 - Dated: 31 Jan 2020 | | ☑ | ☐ |
| | Participant post medical circumcision education checklist - Version: Version 1.0 - Dated: 31 Jan 2020 | | ☑ | ☐ |
| Insurance/Compensation | Hollard - Policy No: SPL/SLFG/000013724 | Valid From: 01 Jan 2020 To: 31 Dec 2020 | ☑ | ☐ |
| Synopsis of Study/Trial Summary | Circumcision Clients in Republic of South Africa | | ☑ | ☐ |
| Other Documentation | Protocol Summary - Dated: 31 Jan 2020 | | ☑ | ☐ |
| | HREC Application Form - Dated: 07 Feb 2020 | | ☑ | ☐ |
| | NHREC Trial Application #9478 - Dated: 05 Feb 2020 | | ☑ | ☐ |
| Relevant Trial Hospital/(s) | The Aurum Institute - The Ridge | | ☑ | ☐ |
| Syndicate and/or Research Unit | The Aurum Institute - The Ridge | | ☑ | ☐ |

UNIVERSITY OF THE WITWATERSRAND, JOHANNESBURG

### INDEPENDENT ETHICS COMMITTEE APPROVAL FORM

| DETAILS OF COMMITTEE | |
|---|---|
| Name | University of the Witwatersrand Human Research Ethics Committee: (Medical) |
| Address | Research Office, Senate House<br>University of the Witwatersrand, 1 Jan Smuts Avenue, BRAAMFONTEIN, Johannesburg, 2000 |

| DETAILS OF MEETING | Yes | No |
|---|---|---|
| Is the Investigator a member of the committee ? | ☐ | ☑ |
| If "Yes" did he/she vote ? | ☐ | ☑ |
| Is the Committee organised and operated according to applicable laws and regulations together with ? | ☑ | ☐ |
| Local GCP requirements ? | ☑ | ☐ |
| ICH GCP requirements ? | ☑ | ☐ |
| FDA GCP requirements ? FWA Registered No. IRB00001223 | ☑ | ☐ |
| Progress reports required either in March and September or six monthly from start of study ? | ☑ | ☐ |

| DECISION ON APPROVAL : is approval given to conduct the trial ? | | Tick As Appropriate |
|---|---|---|
| Yes - with no conditions | | ☐ |
| Remarks : | | |
| Yes - with conditions | | ☑ |
| Specify conditions : | Approved on the following conditions:<br>1. Please submit GCP and Ethics Training Certificates for Ms J Pienaar<br><br>Our expectation is that our requirements are fulfulled once a company has submitted and uploaded relevant approvals onto NHRD. | |
| No | | ☐ |
| Specify reasons | | |

| SIGNATURES | | |
|---|---|---|
| I confirm that the details on this form are correct: | | Date |
| **Name:**<br>**Prof C Penny**<br>**Chair / Deputy Chair of Committee** | **Signature:** [signature] | **29 June 2020** |

**DECLARATION OF INVESTIGATOR/(S)**

To be completed and ONE COPY returned to the Secretariat for the HREC at Wits Health Consortium, 31 Princess of Wales Terrace, Parktown, 2193 or Fax To: 011 274-9281

I/We fully understand the conditions under which I am/we are authorised to carry out and complete the above-mentioned research and I/we agree to ensure full compliance with these conditions. Should any amendment, alteration or departure be contemplated from the research procedure methodology or manner of execution, I/we will communicate with the Chairman of the Human Research Ethics Committee: (Medical) for approval prior to acting on any of the above mentioned proposed amendments, alterations or departures. I am/we are fully aware that any unauthosised amendment, alteration or departure as above will amount to misconduct and may lead to the institution of disciplinary procedures.

Any approval given by the HREC is conditional upon consent being obtained by the Investigator/s from the Superintendent (or equivalent official) of the Hospital, Clinic or Institution in which the research is, in part or full, to take place.

The Chairman may of course at his discretion place the matter before the full Committee.

DATE:__________ SIGNATURE:__________ NAME:__________

PROTOCOL NUMBER **AUR2-8-270 2WT** ETHICS REF.: **200204**

Date Printed 29 June 2020

UNIVERSITY OF THE WITWATERSRAND, JOHANNESBURG

### HUMAN RESEARCH ETHICS COMMITTEE MEMBERS: (MEDICAL) UNIVERSITY OF THE WITWATERSRAND

#### Attendance Register for the Ethics Meeting held on 28 February 2020 from 12:00 - 15:00
#### Venue: EXECUTIVE COMMITTEE ROOM, Ground Floor, Phillip V Tobias Building, Cnr York Road & 29 Princess of Wales Terrace

##### AFFILIATED TO THE UNIVERSITY OF THE WITWATERSRAND

| Surname | Initials | Title | Discipline/s | Academic Qualifications | Gender | Present |
|---|---|---|---|---|---|---|
| Adam | Y | Prof | Obstetrics & Gynaecology | MB BCh; FCOG | F | Present |
| Badal-Faesen | S | Dr | Infectious Diseases/HIV/TB | MB BCh | F | Absent |
| Conradie | FM | Dr | Infectious Diseases/HIV/TB | MB BCh; DTM&H; MSc; Dip HIV Man | F | Absent |
| Cooper | PA | Prof | Paediatrics | MB BCh, PhD, DCH (SA), FCPaeds (SA) | M | Present |
| de Andrade | VM | Dr | Speech Pathology | BA; MA; PhD | M | Present |
| Dhai | A | Prof | Biomedical Ethics | MB ChB; FCOG; LLM; PGDipIntResEthics | F | Present |
| Donde | B | Prof | Radiation Oncology | MB BCh, MMed Rad (T) | M | Present |
| Etheredge | H | Dr | Biomedical Ethics | MSc Med, BA; PhD | F | Present |
| Feldman | C | Prof | Pulmonology | MB BCh, PhD, DSc, FCP (SA), FRCP | M | Absent |
| Gerrand | P | Dr | Social Work | PhD (Social Work) | F | Present |
| Kerr | R | Dr | Human Genetics | BSc, BScHons, MSc, PhD | F | Absent |
| Lownie | MA | Prof | Maxillo-Facial & Oral Surgery | BDS, BA (Hons), DipMFOS, FCMFOS(SA), MEd | F | Absent |
| Menezes | CN | Prof | Internal Medicine | MD, MMed (Int Med), Dip HIV Mang (SA), DTM&H, FCP (SA), Cert ID (SA), PhD | M | Absent |
| Mokhachane | M | Dr | Clinical Medicine | MB BCh, FCP (Paeds) SA, MMed, Neonatology (SA) | F | Present |
| Naidoo | S | Prof | Public Health | MB BCh, DMTH, DHSM, DOH, MMED | M | Absent |
| Naran | NH | Dr | Chemical Pathology | PhD | M | Absent |
| Penny | C | Prof | Internal Medicine | BSc Hons, PhD | M | Present |
| Ross | M | Prof | Public Health | MB ChB, FFCH(SA), FOM(UK) | F | Absent |
| Sanne | IM | Prof | Infectious Diseases/HIV/TB | MB BCh, FCP (SA), DTM&H; MMed & PhD | M | Absent |
| Smith | C | Prof | Psychiatry | BA, BA (Hons), M.A (Clin.Psych), PhD | F | Present |
| Stewart | A | Prof | Physiotherapy | BSc (Physio), MSc, PhD, DPE | F | Absent |
| Szabo | CP | Prof | Psychiatry | MB BCh, MMed, MScMed, PhD; FCPsych(SA) | M | Absent |
| Thom | RGM | Prof | Psychiatry | MB ChB, DCH, FCPsych, PhD | F | Absent |
| Tsotsi | NM | Dr | | BDS; MPH; MSc Med; PGDipInt ResEthics | F | Absent |
| Velaphi | S | Prof | Paediatrics | MB BCh, FCPaeds, MMed, PhD | M | Present |
| Wadee | R | Dr | Anatomical Pathology | MBBCh, FCPath, MMed, PhD | F | Present |
| Warria | A | Dr | Social Work | D Litt et Phil (SWK) | F | Absent |
| Willem | P | Dr | Human Genetics | MD, PhD | F | Absent |
| Woodiwiss | AJ | Prof | Cardiovascular Pathophysiology | BSc Physiotherapy, BSc, MSc, PhD | F | Absent |

##### NOT AFFILIATED TO THE UNIVERSITY OF THE WITWATERSRAND

| Surname | Initials | Title | Discipline/s | Academic Qualifications | Gender | Present |
|---|---|---|---|---|---|---|
| Barnabas | N | Ms | Civil Society Liaison Officer | Community Liaison Manager | F | Absent |
| Burns | I | Mr | Community Representative | MA (Hons) | | Present |
| Egan | A | Father | Theology | BA (Hons), MA, MDiv, STL, PhD | M | Absent |
| Guidozzi | Y | Adv | Lawyer | BSc (Nurs), LLB, MBA | F | Absent |
| Ikalafeng | B | Dr | Governance | BSc (Hons), MSc, PhD | F | Absent |

| | | | | | | |
|---|---|---|---|---|---|---|
| Langley | G | Dr | Nursing | MSc (Nursing), PhD, MPhil (Ethics); BA (Hons) Theol | F | Present |
| Paruk | F | Prof | Anaesthesia | MB ChB, FCOG(SA), Crit Care(SA), PhD | F | Absent |
| Peter | JR | Adv | Lawyer | BCom; LLB; LLM | M | Absent |
| Van Gelderen | CJ | Prof | Obstetrics & Gynaecology | MB BCh, FRCOG, FCOG(SA) | M | Present |

**RETIRED MEMBER OCCASIONALLY CO-OPTED FOR SURGICAL OPINION ON A PROJECT**

| | | | | | |
|---|---|---|---|---|---|
| Oettle | GJ | Prof | Surgery | BSc (Hons), MB BCh, FRCS | M |

**Note 1:** This committee has been in continuous operation since October 1966

**Note 2:** The large committee size is to ensure a good attendance at meetings

**Note 3:** A Quorum is 12 members according to the 33% of members on a committee with more than 15 members as required by SA National Guidelines (ref 2 below)

**This is to certify that the Human Research Ethics Committee: (Medical) of the University of the Witwatersrand operates according to the following guidelines of good clinical practice:**

1. **ICH Harmonised Tripartite Guideline for Good Clinical Practice.**
2. **SA National Department of Health 2006 Guidelines for Good Practice in the Conduct of Clinical Trials with Human Participants in South Africa (2006).**
3. **Declaration of Helsinki 2013.**

**The Committee's United States Federal Wide Assurance details are:**

1. **Country code SF.**
2. **FWA Number: FWA00000715.**
3. **University of the Witwatersrand: IORG0000862.**
4. **Human Research Ethics Committee: (Medical): IRB00001223.**
