## Supplementary material for "Strengthening evidence for text-based telehealth in post-operative care: A pragmatic study of the reach and effectiveness of two-way, text-based follow-up after voluntary medical male circumcision in South Africa": U Washington Ethical Approval

UNIVERSITY of WASHINGTON
HUMAN SUBJECTS DIVISION

### IRB APPROVAL OF APPLICATION

April 6, 2020

Dear Caryl B Feldacker:

On 4/6/2020, University of Washington IRB Committee J reviewed the following application:

| | |
|---|---|
| Type of Review: | Initial Study |
| Title of Study: | Expanding and Scaling Two-way Texting to Reduce Unnecessary Follow-Up and Improve Adverse Event Identification Among Voluntary Medical Male Circumcision Clients in the Republic of South Africa |
| Investigator: | Caryl B Feldacker |
| IRB ID: | STUDY00009703 |
| Funding: | Name: National Institutes of Health (NIH), Grant Office ID: A151492, Funding Source ID: 12992084 |
| IND, IDE, or HDE: | None |

**IRB Approval**

Under FWA #00006878, the IRB approved your activity.

- **Depending on the nature of your study, you may need to obtain other approvals or permissions to conduct your research. For example, you might need to apply for access to data or specimens (e.g., to obtain UW student data). Or, you might need to obtain permission from facilities managers to approach possible subjects or conduct research procedures in the facilities (e.g., Seattle School District; the Harborview Emergency Department).**
- Your application qualified for expedited review ("minimal risk"; Categories 5, 6, and 7).
- Under the Revised Common Rule this IRB approval is valid until study completion. In other words, there is no expiration date and you are not required to submit Continuing Review Reports to maintain your approval. However, you are still required to (1) obtain IRB approval before making any changes (modifications) to your research, and (2) provide the IRB with any Reportable New Information such as breaches of confidentiality or unanticipated problems.
- This approval applies only to the activities described in your application (including any references to specific grant sections). It does not include other activities that may be described in your grant or contract.
- Your study automatically has a Certificate of Confidentiality (CoC), because you have NIH funding. A description of the CoC protections and responsibilities has been placed in your study's Documents section.
- If you plan to continue data collection past the expiration of your NIH funding and the CoC, contact the Human Subjects Division prior to the end of your funding. We will help you determine whether you need to apply for a CoC extension.

Determinations, waivers, and regulations

The IRB made the determinations and waivers listed in the table below. Note that any granted waivers of consent or parent permission do not override a subject's refusal to provide broad consent.

| Requirement | Determination or Waiver |
| --- | --- |
| Consent | Waived for clinicians observed as part of the time and motion portion of 2WT populations study procedures. |

Location of documents

Use the consent forms that were approved and stamped by the IRB. They can be downloaded from the Final column under the **Documents tab** in Zipline.

In addition, HSD has uploaded the following documents to the **Documents tab** in Zipline:

- Certificate of Confidentiality Acknowledgement Letter

Thank you for your commitment to ethical and responsible research. We wish you great success!

Sincerely,

Jeff Love, IRB Administrator
206-543-2921,
